# Intravenous thrombolysis at primary stroke centers versus comprehensive stroke centers: Analysis from the AcT trial

**DOI:** 10.1101/2025.03.05.25323461

**Authors:** Diego Gutiérrez, Katrina Ignacio, Aleksander Tkach, Herbert Manosalva, Aravind Ganesh, MacKenzie Horn, Houman Khosravani, Jai Shankar, Fouzi Bala, Mohamed A. Tarek, Gary Hunter, Ayoola Ademola, Dar Dowlatshahi, Andrew M. Demchuk, Richard H Swartz, Luciana Catanese, Tolulope T Sajobi, Mohammed A. Almekhlafi, Bijoy K. Menon, Nishita Singh

**Affiliations:** Calgary Stroke Program, Department of Clinical Neurosciences, Cumming School of Medicine, University of Calgary, Calgary, AB, Canada; Department of Neurology, Pontifical Catholic University of Chile, Santiago, Chile; Kelowna General Hospital, Kelowna, BC, Canada; Medicine Hat Regional Hospital, AB, Canada; Department of Clinical Neurosciences, Cumming School of Medicine, University of Calgary, Calgary, AB, Canada; Department of Radiology, University of Calgary, Calgary, AB, Canada; Department of Community Health Sciences, Cumming School of Medicine, University of Calgary, Calgary, AB, Canada; Hotchkiss Brain Institute, Cumming School of Medicine, University of Calgary, Calgary, AB, Canada; Hurvitz Brain Sciences Program, Sunnybrook Health Sciences Centre, University of Toronto, Toronto, ON, Canada; Department of Internal Medicine-Neurology Division, Rady Faculty of Health Sciences, University of Manitoba, Winnipeg, MB, Canada; Diagnostic and Interventional Neuroradiology Department, University Hospital of Tours, Tours, France; Hamilton Health Sciences Centre, McMaster University, Hamilton, ON, Canada; University of Saskatchewan, Saskatoon, SK, Canada; Department of Medicine, University of Ottawa and the Ottawa Hospital Research Institute, Ottawa, ON, Canada; Division of Neurology, Sunnybrook Health Sciences Centre, University of Toronto, ON, Canada

**Keywords:** Intravenous thrombolysis, tenecteplase, primary stroke centers, workflow

## Abstract

**Introduction:** Fast delivery of intravenous thrombolysis (IVT) and transportation to a comprehensive stroke center (CSC) is paramount in primary stroke centers (PSCs), to achieve effective reperfusion. We investigated clinical outcomes and workflow times of patients treated with IVT at PSCs.

**Methodology:** This is a secondary analysis of the AcT trial, a multicenter, phase-3, randomized, controlled, noninferiority trial comparing tenecteplase with alteplase in patients with acute ischemic stroke within 4.5 hours of onset. We compared baseline characteristics, imaging and clinical outcomes at 90 days, and workflow times between PSCs and CSCs.

**Results:** Of 1577 patients enrolled in the trial, 99 (6.27%) were treated at PSCs and 1,478 (93.72%) at CSCs. Both groups had similar age (median 72 [64 - 82] versus 74 [63 - 83] years), proportion of females (42.42% versus 48.24%), baseline stroke severity (median National Institute of Health Stroke Scale 9 [6 - 16] versus 10 [6 - 16.5] points) and presence of large vessel occlusion (24.24% versus 24.70%). The proportion of patients achieving excellent functional outcome at 90 days was significantly higher in PSCs compared to CSCs (mRS 0-1: 48.48% versus 35.01%, adjusted IRR, 1.42 [CI 95%, 1.04 - 1.95]), without differences in safety outcomes. Patients treated at PSCs had longer onset-to-needle (median, 139 [100 - 190] versus 128 [94 - 185] minutes, p 0.026) and door-to-needle times (median, 56.5 [42 - 70] versus 35 [27 - 47] minutes, p <0.001). In the 24 patients transferred from PSCs to CSCs, those who received tenecteplase had lower needle-to-puncture times compared to those who received alteplase (median, 35.5 [21 - 58] versus 52 [18 - 74] minutes, p <0.001).

**Conclusions:** Despite less efficient workflows for IVT and similarities in baseline characteristics, the proportion of patients treated at PSCs who had excellent functional outcomes was higher compared to CSCs. Patients who received tenecteplase prior to CSC transfer had more efficient time metrics when compared to those who received alteplase.

## Introduction

Intravenous thrombolysis (IVT) is a specific, effective, time - dependent reperfusion therapy for acute ischemic stroke. Each 15-minute decrease in treatment delay is estimated to provide an average of one month of additional disability - free life.^1^ This increased chance of achieving better clinical outcomes with faster administration has not only been observed with alteplase but also with tenecteplase, demonstrating a 1.8% and 0.2% increase in the probability of achieving excellent functional outcome at 90 days with a 30-minute reduction in the onset-to-needle time, and with every 10-minute reduction in door-to-needle time, respectively.^2^ Fast delivery of this treatment can be accomplished not only in comprehensive stroke centers (CSCs) that offer more advanced therapies (i.e., endovascular thrombectomy (EVT), decompressive craniectomy, intensive care unit), but also in centers that have the capability only to offer urgent neuroimaging and to administer IVT.^3^ These primary stroke centers (PSCs) usually are located in suburban and rural areas, and they can be the closest hospital able to offer prompt diagnosis, urgent neuroimaging and IVT treatment to their catchment area. Considering the time-sensitive nature of stroke therapies, it is recommended that patients with suspected acute stroke be transported to the nearest stroke center.^4^

A definite pathway for patients with suspected large vessel occlusion (LVO) regarding transportation to PSCs or CSCs is still in question. Attempts to model a decision-making process for patients with LVO have been made taking into consideration several challenges including regional geography, transportation times, and treatment times at these facility types.^5^ In addition, it is hypothesized that tenecteplase with its ease of use could contribute to faster IVT delivery in any center and decrease transfer times from PCSs to CSCs, a critical aspect especially in patients eligible to receive EVT.^6^ The pharmacokinetic and pharmacodynamic profile of tenecteplase is better than alteplase, with longer half-life, less binding to plasminogen activator inhibitor-1 and higher fibrin specificity.^7^

Treatment of acute ischemic stroke is highly time-sensitive, where PSCs play a key role in treating patients. It is therefore of utmost importance to efficiently transfer patients with suspected LVOs from PSCs to CSCs. In this secondary analysis of the AcT (Alteplase Compared to Tenecteplase) registry-linked randomized controlled trial (RCT) trial, we sought to investigate the clinical outcomes and workflow times of patients treated with IVT in PSCs. In addition, we evaluated key metrics in patients transported from PSCs to CSCs for potential EVT enrolled in this trial.^8^

## Methods

### Study design and population

This is a subgroup analysis from the AcT trial.^9^ AcT was a phase 3, multicenter, prospective, pragmatic randomized controlled trial that assessed the non-inferiority of tenecteplase compared to alteplase for IVT in eligible patients aged ≥ 18 years with an acute ischemic stroke within 4.5 hours. A total of 22 centers participated in the trial, of which five were PSCs and the rest were CSCs. PSCs were defined as hospitals with resources and processes to offer IVT to patients with an acute ischemic stroke, but without the capability to offer EVT. CSCs were defined as centers with capability to offer IVT and EVT.

### Efficacy and Safety Outcomes

The primary outcome was the proportion of patients with excellent functional outcome defined as having an mRS 0 - 1 at 90 days to 120 days after randomization. We assessed the following secondary outcomes: mRS 0 - 2 at 90 to 120 days, actual mRS score achieved at 90 to 120 days, return to baseline function (defined as return to pre-stroke function), EQ-VAS at 90 to 120 days, Euro QOL (EQ5D5L) and EQ5D index, length of hospital stay, length of hospital stay for more than seven days, EVT utilization rates, eTICI score ≥2b and rAOL score of ≥2b on initial angiography for those who underwent EVT.

Additionally, we analyzed the following safety outcomes: mortality at 90 days, symptomatic intracerebral hemorrhage (sICH) at 24 hours, defined as any intracerebral hemorrhage that was temporally related to, and directly responsible for worsening of the patient’s neurological condition, other intracranial bleeding (subarachnoid hemorrhage, subdural hemorrhage and intraventricular hemorrhage), peripheral bleeding requiring blood transfusions, any hemorrhage, orolingual angioedema and other serious adverse events.

### Workflow time metrics

The following data on prehospital and in-hospital workflow times were collected from linked registries (QuiCR for Alberta and OPTIMISE for all other Canadian provinces except for Alberta): For both PSC and CSC: onset-to-door time (time of stroke symptom onset to patient arrival at the emergency department [ED]), onset-to-randomization time (time of stroke symptom onset to randomization), onset-to-needle time (time of stroke symptom onset to IVT administration), door-CT-time (time of emergency department arrival to first slice of CT imaging); door-to-needle time (time of ED arrival to IVT administration), and for CSC only: onset-to-puncture time (time of stroke symptom onset to groin puncture for EVT), door-to-puncture time (time of ED arrival to groin puncture time for EVT); door-to-reperfusion time (time of ED arrival to reperfusion time for EVT) and puncture-to-first-reperfusion time (time of groin puncture to reperfusion time for EVT). First reperfusion time is defined as the first time recanalization status was achieved during the procedure.

Additionally, for patients transferred from PSCs to CSCs for potential EVT, the following data was collected: onset-to-door time and onset-to-CT time at PSCs, CT-to-puncture time, onset-to-needle time, needle-to-puncture time (time of IVT administration to groin puncture time for EVT), eTICI score of ≥2b/3 on initial angiography run for EVT, puncture-to-successful reperfusion time and needle-to-first reperfusion time (time from IVT administration to time recanalization status achieved).

### Statistical Analysis

Descriptive analysis was performed to compare baseline demographic and clinical characteristics of patients enrolled at PSCs and those enrolled at CSCs. Baseline characteristics were summarized using frequencies and proportions for categorical variables, mean with standard deviation (for normally distributed continuous variables) and medians with interquartile ranges (for non-normally distributed continuous variables). Mixed-effects Poisson regression analysis was conducted to determine the association between center type (PSC vs CSC) and the respective primary, secondary and safety outcomes. Age, sex, baseline stroke severity (NIHSS), location of intracranial occlusion, door-to-needle time and thrombolytic drug given were included as fixed effects variables and site was included as a random effects variable. For models assessing workflow times as an outcome, mixed-effects Poisson regression analysis was used adjusting for age, sex, NIHSS, drug type and location of occlusion as fixed effects variables and participating site as random effects variable.

We report effect estimates as rate ratios (adjusted and unadjusted). A p-value of <0.05 was considered significant. Similar unadjusted and adjusted analysis was done comparing differences in outcomes between alteplase and tenecteplase in patients who presented at CSC and PSC respectively. All analysis was considered exploratory. In addition, we also conducted pre-specified analysis to assess heterogeneity in treatment effects in the intention-to-treat population for primary outcome (mRS 0 - 1 at 90 to 120 days) and key safety outcomes by type of enrolling hospital (PSCs vs. CSCs). Evidence of a treatment-by-sub-group variable interaction was tested by including a multiplicative interaction term (thrombolytic type*center type) in the mixed effects logistic regression model adjusted for age, sex, baseline stroke severity, and stroke onset-to-needle time as fixed effects, and site, and registry (QuiCR vs. OPTIMISE) as random effects. Statistical significance for each subgroup analysis was considered exploratory and conducted at alpha <0.05.

The variables chosen for the adjusted analysis were selected to account for known confounders influencing outcomes, including demographic, clinical, and treatment-related factors. In contrast, the pre-specified analysis focused on assessing treatment effect heterogeneity by center type, prioritizing key modifiers of treatment response while avoiding over-adjustment. Stata v17.0 software was used for all analyses.

### Standard Protocol Approvals, Registrations, and Patient Consents

Approval form the ethical standards committee was received to conduct this secondary analysis.

### Data availability

Anonymized data not published within this article will be made available by request from any qualified investigator. Please send data access requests to. Such requests must be approved by the respective ethics boards and appropriate data custodians.

## Results

### Baseline characteristics

Of 1577 participants enrolled in the AcT trial, 99 (6.27%) were enrolled at PSCs (56: tenecteplase, 43: alteplase) and 1478 (93.72%) (750 tenecteplase, 728: alteplase) at CSCs. Baseline demographic, clinical and radiological characteristics were similar between the two groups and by thrombolytic type (Table 1 and Supplemental Table 1).

**Table 1.** Baseline demographic, clinical and radiological characteristics of patients treated in PSCs and CSCs.

|  | **PSC**<br>(n = 99) | **CSC**<br>(n = 1,478) | **p value** |
| --- | --- | --- | --- |
| Age, years (median, IQR) | 72 (64 - 82) | 74 (63 - 83) | 0.66 |
| Female (n, %) | 42 (42.4) | 713 (48.2) | 0.26 |
| Baseline NIHSS score (median, IQR) | 9 (6 - 16) | 10 (6 - 16.5) | 0.83 |
| Baseline NIHSS score categories (n, %)* |  |  |  |
| < 8 | 45 (45.9) | 574 (39.0) | 0.28 |
| 8 - 15 | 25 (25.5) | 479 (32.5) |  |
| > 15 | 28 (28.8) | 419 (28.5) |  |
| ASPECTS (median, IQR) | 9 (8 - 10) | 9 (8 - 10) | 0.34 |
| Occlusion site on baseline CT angiography (n, %) |  |  |  |
| Intracranial internal carotid artery | 6 (6.1) | 129 (8.7) | 0.36 |
| M1 segment MCA | 17 (17.2) | 222 (15.0) | 0.56 |
| M2 segment MCA | 17 (17.2) | 300 (20.3) | 0.45 |
| Other distal occlusions (MCA, ACA, PCA) | 19 (19.2) | 292 (19.8) | 0.89 |
| Vertebrobasilar circulation | 1 (1.0) | 63 (4.3) | 0.11 |
| Cervical internal carotid artery | 1 (1.0) | 25 (1.7) | 0.61 |
| No visible occlusions† | 39 (40.6) | 474 (32.4) | 0.10 |
| Presence of large vessel occlusion (LVO) on baseline CTA (n, %) | 24 (24.2) | 365 (24.7) | 0.92 |
| Source registry (n, %) |  |  |  |
| QuiCR | 36 (36.4) | 652 (44.1) | 0.13 |
| OPTIMISE | 63 (63.6) | 826 (55.9) |  |

IQR, interquartile range; NIHSS, National Institutes of Health Stroke Scale; ASPECTS, Alberta Stroke Program Early CT Score; ICA, internal carotid artery; MCA, middle cerebral artery; ACA, anterior cerebral artery; PCA, posterior cerebral artery; LVO, large vessel occlusion.
\*In 1570 patients. †In 1558 patients. Analysis: Mann-Whitney (age, NIHSS). Chi2.

### Outcomes

The proportion of excellent functional outcome (mRS 0-1) at 90 to 120 days was significantly higher for patients treated at PSCs compared to CSCs (48.48% versus 35.01%; adjusted IRR, 1.42 [95% CI, 1.04 - 1.95, p = 0.026]). Ordinal mRS and return to baseline function were statistically different between the PSC and CSC group, showing lower mRS scores (median, 2 (0 - 3) vs 2 (1 - 4) points; adjusted IRR, 0.48 [95% CI 0.32 - 0.71) and higher proportion of patients with return to baseline pre-stroke function among patients enrolled at PSCs (48.31% versus 27.47%; adjusted IRR 1.77 (95% CI 0.09 - 2.87) (Figure 1). Likewise, EQ-VAS score at 90 days was significantly higher in patients treated at PSCs compared to CSCs [median (IQR), 80 (53 - 90) versus 75 (58 - 90) points; adjusted IRR 0.62 [95% CI 0.45 - 0.85]. Rate of EVT utilization, successful reperfusion on initial angiographic run in patients undergoing EVT was similar (Table 2 and Supplemental Table 2.)

**Table 2.** Efficacy and safety outcomes in patients treated in PSCs compared to CSCs.

|  | PSC<br>(n = 99) | CSC<br>(n = 1478) | Unadjusted IRR (95% CI) | Adjusted IRR (95% CI) |
| --- | --- | --- | --- | --- |
| **Primary outcome** |  |  |  |  |
| mRS score 0 - 1 at 90 - 120 days (n, %)* | 48 (48.48) | 514 (35.01) | 1.38 (1.03 - 1.86) | 1.42 (1.04 - 1.95) |
| **Secondary outcomes** |  |  |  |  |
| mRS score 0 - 2 at 90 - 120 days (n, %)* | 67 (67.68) | 810 (55.18) | 1.23 (0.96 - 1.57) | 1.26 (0.97 - 1.64) |
| Actual mRS score at 90 - 120 days (median, IQR) | 2 (0 - 3) | 2 (1 - 4) | 0.53 (0.37 - 0.78) §§ | 0.48 (0.32 - 0.71) §§ |
| Return to baseline function (n, %)** | 43 (48.31) | 375 (27.47) | 1.76 (1.28 - 2.41) | 1.77 (0.09 - 2.87) |
| EQ-VAS at 90 - 120 days (median, IQR)† | 80 (53 - 90) | 75 (58 - 90) | 0.95 (0.93 - 0.97) | 0.62 (0.45 - 0.85) |
| EQ-5D index (median, IQR) | 0.68 (0.46-0.76) | 0.61 (0.35-0.74) | 0.79 (0.54-1.15) | 0.70 (0.41-1.21) |
| Length of hospital stay (days), median (IQR)§ | 3 (1 - 9) | 5 (3 - 11) | 0.88 (0.83 - 0.95) | 1.02 (0.71 - 1.47) |
| Length of hospital stay > 7 days, n (%) | 33 (33.33) | 606 (41.00) | 0.81 (0.57 - 1.15) | 0.82 (0.51 - 1.33) |
| EVT utilization, n (%) | 24 (24.24) | 482 (32.61) | 0.74 (0.49 - 1.12) | 0.85 (0.55 - 1.30) |
| eTICI score of ≥2b on initial angiography of EVT (n, %)¶ | 1 (4.17) | 51 (10.67) | 0.39 (0.05 - 2.83) | 0.43 (0.59 - 3.20) |
| rAOL score of ≥2b on initial angiography of EVT (median, IQR)‡ | 2 (8.33) | 86 (18.07) | 0.46 (0.11 - 1.87) | 0.58 (0.14 - 2.43) |
| **Safety outcomes** |  |  |  |  |
| Mortality at 90 days (n, %)** | 12 (12.12) | 229 (15.60) | 0.78 (0.43 - 1.39) | 0.81 (0.45 - 1.47) |
| Symptomatic intracerebral hemorrhage at 24 h (n, %)†† | 3 (3.06) | 40 (2.72) | 0.61 (0.15 - 2.51) | 0.62 (0.15 - 2.58) |
| Other intracranial bleeding (n, %)**** |  |  |  |  |
| Subarachnoidal hemorrhage | 3 (3.06) | 102 (7.00) | 0.44 (0.14 - 1.38) | 0.44 (0.13 - 1.44) |
| Subdural hemorrhage | 0 (0.00) | 7 (0.48) | - | - |
| Intraventricular hemorrhage | 2 (.04) | 39 (2.67) | 0.76 (0.18 - 3.16) | 0.97 (0.22 - 4.20) |
| Peripheral bleeding requiring blood transfusions (n, %)†† | 0 (0.00) | 1 (0.07) | - | - |
| Any hemorrhage (n, %)***** | 21 (21.43) | 291 (19.96) | 1.07 (0.69 - 1.67) | 1.26 (0.72 - 2.22) |
| Orolingual angioedema (n, %)†† | 1 (1.02) | 8 (0.54) | 0.93 (0.12 - 7.04) | 0.98 (0.11 - 8.53) |
| Other serious adverse events (n, %)†† | 8 (8.16) | 124 (8.42) | 0.30 (0.10 - 0.94) | 0.44 (0.10 - 1.93) |

mRS, modified Rankin Scale; EQ-VAS, EuroQol-visual analogue scales; EVT, endovascular thrombectomy; rAOL, revised Arterial Occlusion scale; eTICI, extended Thrombolysis in Cerebral Infarction, NIHSS, National Institutes of Health Stroke Scale.
\*In 1567 patients. †In 1333 patients. §In 1481 patients. ¶In 506 patients. ‡In 500 patients. \*\*In 1567 patients. ††In 1570 patients. \*\*\*89 patients. \*\*\*\* in 1556 patients.
Analysis: Poisson. §§ OR (ologit). Adjusted for age, sex, baseline NIHSS, intracranial occlusion, drug type and door-to-needle time.

**Figure 1.**
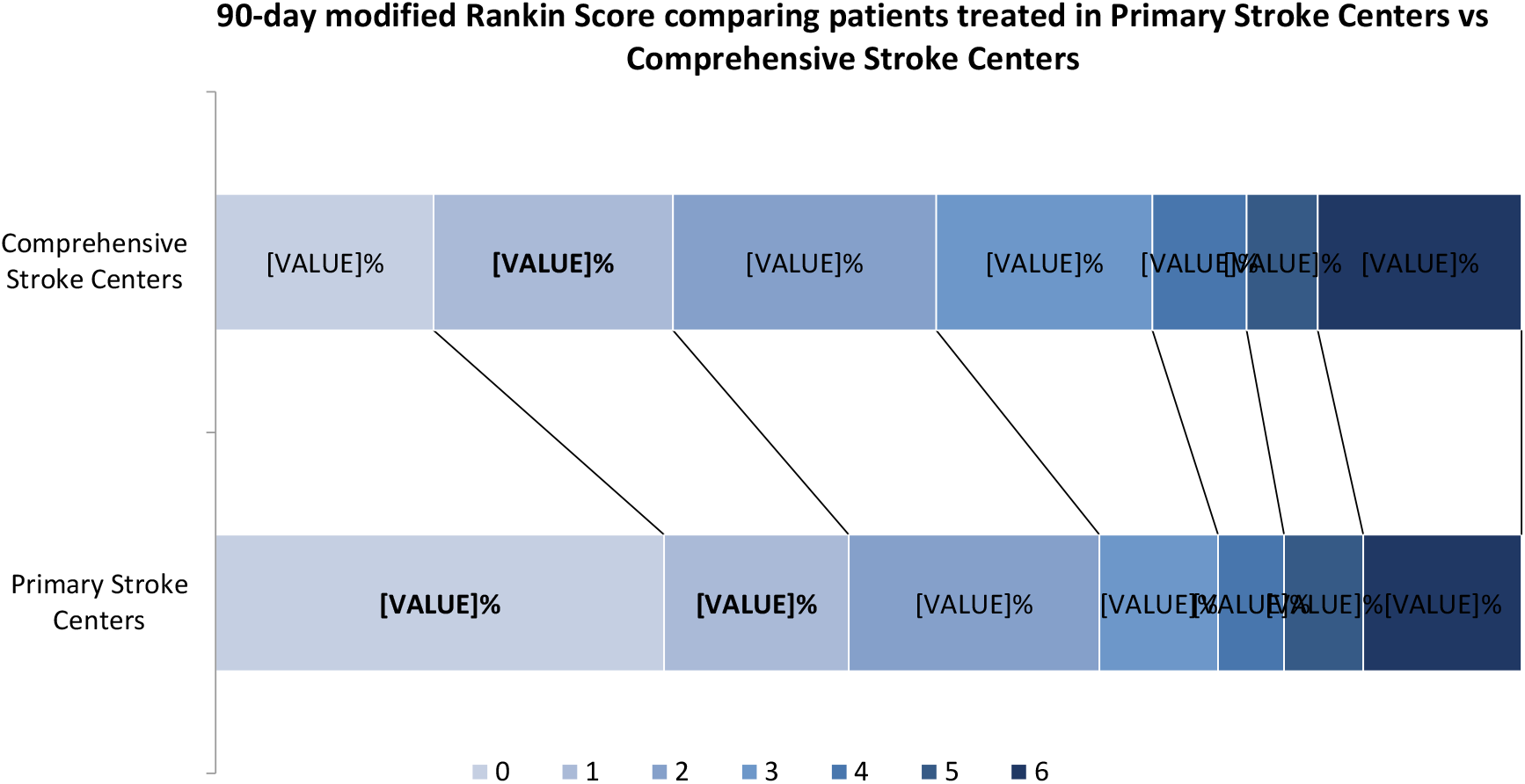
90-day mRS comparing patients treated in PSCs vs CSCs.

All safety outcomes including mortality at 90 days, symptomatic intracerebral hemorrhage at 24 hours, peripheral bleeding requiring transfusion, orolingual angioedema and other serious adverse events were similar between the two groups (Table 2, supplemental Table 2 and 3).

There was no difference in primary outcome between patients treated with tenecteplase versus alteplase at PSCs (50% versus 46.51%; adjusted IRR 1.01 [95% CI 0.58 - 1.75) and at CSCs (35.92% versus 34.07%; adjusted IRR 1.08 [95% CI 0.95 - 1.23). Likewise, safety outcomes were similar stratifying by thrombolytic type in PSCs and CSCs (Supplemental Tables 2 and 3).

### Workflow metrics

Onset-to-needle (median (IQR) 139 (100 - 190) versus 128 minutes (94 - 185); adjusted IRR 1.11 [95% CI 1.01 - 1.23]) and door-to-needle times (median (IQR) 56.5 (42 - 70) versus 35 minutes (27 - 47)); adjusted IRR 1.57 [95% CI 1.27 - 1.96]) were significantly longer at PSCs compared to CSCs (Figure 2). There was no difference between PSC and CSC for other workflow metrics such as onset-to-door, onset-to-CT, door-to-CT and onset-to-randomization times (Table 3).

**Table 3.** Workflow times in patients arriving at PSCs and CSCs.

|  | **PSC**<br>(n = 99) | **CSC**<br>(n = 1478) | **Unadjusted IRR**<br>**(95% CI)** | **Adjusted IRR (95% CI)** |
| --- | --- | --- | --- | --- |
| Onset-to-door time (min [median, IQR])* | 67 (40 - 117) | 83 (55 - 140) | 0.85 (0.84-0.87) | 0.93 (0.80-1.09) |
| Onset-to-CT (min [median, IQR]) | 93.5 (62 - 135) | 102 (72 - 158) | 0.91 (0.89-0.93) | 0.97 (0.86-1.09) |
| Onset to randomization (min [median, IQR])<sup>†</sup> | 126.5 (85 - 181) | 122 (87 - 179) | 1.01 (0.99-1.03) | 1.08 (0.98-1.19) |
| Onset-to-needle time (min [median, IQR])<sup>§</sup> | 139 (100 - 190) | 128 (94 - 185) | 1.05 (1.03-1.07) | 1.11 (1.01-1.23) |
| Door-to-CT time (min [median, IQR])** | 20 (11 - 27) | 15 (12 - 20) | 1.20 (1.16-1.26) | 1.21 (0.96-1.53) |
| Door-to-needle time (min [median, IQR])<sup>††</sup> | 56.5 (42 - 70) | 35 (27 - 47) | 1.55 (1.51-1.60) | 1.57 (1.27-1.96) |
| Onset-to-puncture time, in patients undergoing EVT (min [median, IQR])<sup>‡</sup> | 169 (121.5 - 249.5) | 156 (122 - 219) | 0.99 (0.96-1.02) | 1.46 (1.15-1.84) |
| CT-to-puncture time (min [median, IQR]) | 74.5 (55 - 94.5) | 58 (42 - 84) | 1.19 (1.14-1.24) | 2.08 (1.35-3.22) |
| Door-to-puncture time, in patients undergoing EVT (min [median, IQR])<sup>§§</sup> | 97 (75.5 - 130) | 74 (57 - 103) | 0.84 (0.81-0.87) | 1.86 (0.90-3.84) |
| Door-to-reperfusion time, in patients undergoing EVT (min [median, IQR])<sup>¶¶</sup> | 147.5 (97 - 202) | 113 (87 - 150) | 1.15 (1.11-1.20) | 1.86 (1.26-2.75) |
| Puncture-to-first-reperfusion, in patients undergoing EVT (min [median, IQR])*** | 24.5 (15.5 - 49) | 29 (18 - 46) | 1.04 (0.97-1.11) | 1.48 (0.92-2.38) |

EVT, endovascular thrombectomy; CI, confidence interval; IQR, interquartile range; NIHSS, National Institutes of Health Stroke Scale. Onset: stroke symptom onset. Door: hospital arrival. Needle: intravenous thrombolysis start. Puncture: groin puncture, EVT start.
\*In 1562 patients. <sup>†</sup>In 1571 patients. <sup>§</sup>In 1563 patients. <sup>¶</sup>In 505 patients. \*\*In 1527 patients. <sup>††</sup>In 1522 patients. <sup>§§</sup>In 503 patients. <sup>¶¶</sup>In 437 patients. \*\*\*In 446 patients.
Analysis: Poisson. Adjusted for age, sex, baseline NIHSS, intracranial occlusion and drug type.

**Figure 2.**
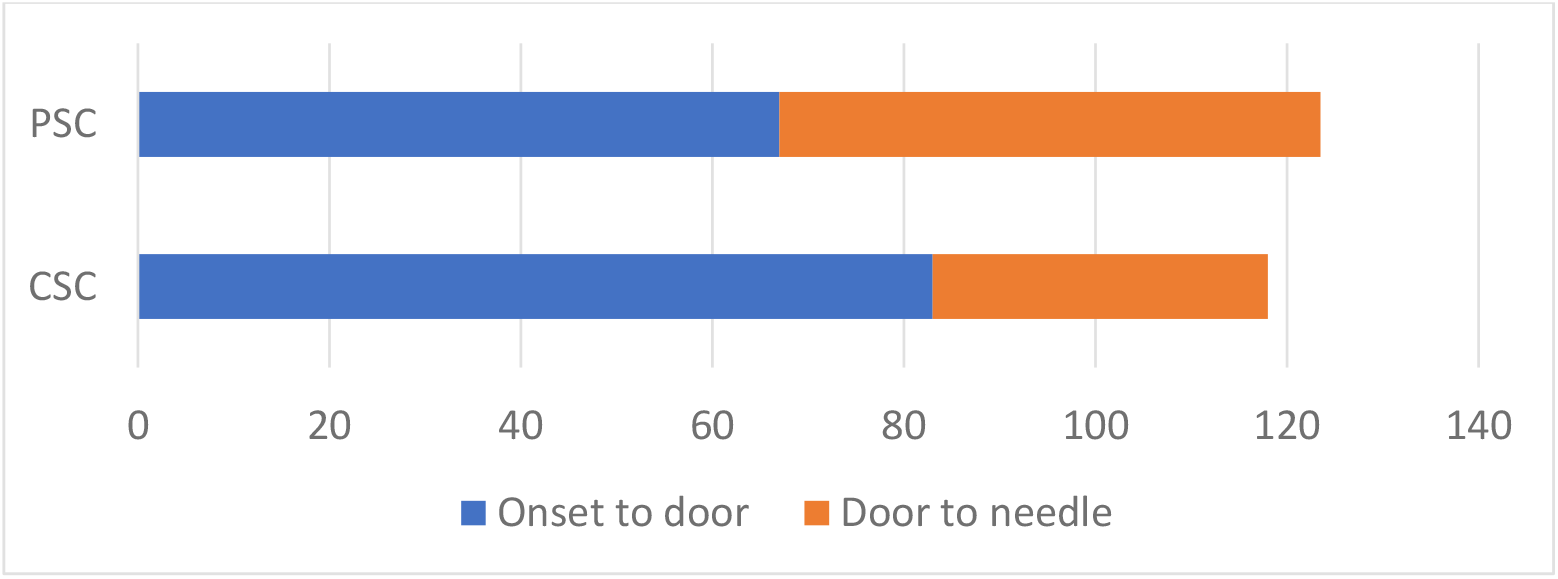
Onset-to-door and door-to-needle times in patients treated in PSCs and CSCs. PSC: Primary Stroke Centers. CSC: Comprehensive Stroke Centers. Values correspond to median. Numbers represent minutes.

For patients who underwent EVT, presentation at a PSC was associated with significantly longer onset-to-puncture (median, 169 versus 156 minutes; adjusted IRR, 1.46 [95% CI, 1.15 - 1.84]), initial CT (at PSC)-to-puncture (median 74.5 versus 58 minutes; adjusted IRR, 2.08 [95% CI, 1.35 - 3.22,) and door-to-reperfusion times (median 147.5 versus 113 minutes; adjusted IRR, 1.86 [95% CI, 1.26 - 2.75) as compared to those presenting at CSC. Puncture-to-first reperfusion time (median, 24.5 versus 29 minutes; adjusted IRR, 1.86 (95% CI, 1.26 - 2.75]) was similar in both groups. (Supplemental table 4).

The workflow times comparing patients who received tenecteplase vs alteplase for the subgroup of patients transferred from PSCs to CSCs are shown in Table 4. There were 24 patients in total who were transferred for EVT, 14 of whom received tenecteplase. The following time metrics were significantly lower in patients that received tenecteplase compared to alteplase: CT-to-puncture time (median, 60.5 versus 92.5 minutes; adjusted IRR, 0.77 [95% CI, 0.69 - 0.85]); needle-to-puncture time (median, 35.5 versus 52 minutes; adjusted IRR, 0.61 [95% CI, 0.54 - 0.69]), and needle-to-reperfusion time (median, 82.5 versus 116 minutes; adjusted IRR: 0.80 [95% CI, 0.72 - 0.89) (Figure 3). Meanwhile, groin-to-reperfusion time was similar between the two groups (median, 29.5 versus 23 minutes; adjusted IRR: 0.94 [95% CI, 0.78 - 1.12]).

**Table 4:** Workflow times in patients transferred from PSCs to CSCs for potential EVT, by thrombolytic type.

|  | **Tenecteplase**<br>**(n = 14)** | **Alteplase**<br>**(n = 10)** | **IRR (95% CI)** | **Adjusted IRR (95% CI)** |
| --- | --- | --- | --- | --- |
| Time from onset to PSC door (median, IQR) | 66 (41 - 83) [n=14] | 53 (34 - 67) [n=10] | 1.16 (1.06 - 1.29) | 0.96 (0.85 - 1.08) |
| Time from onset to CT at PSC (median, IQR) | 82.5 (61 - 116) [n=14] | 76 (58 - 98) [n=10] | 1.08 (0.99 - 1.18) | 0.89 (0.80 - 0.98) |
| Time from onset to needle (median, IQR) | 111 (90 - 150) [n=14] | 104 (82 - 175) [n=10] | 1.03 (0.96 - 1.11) | 0.92 (0.84 - 1.00) |
| Time from CT at PSC to puncture (median, IQR) | 60.5 (56 - 92) [n=14] | 92.5 (51 - 95) [n=10] | 0.88 (0.81 - 0.95) | 0.77 (0.69 - 0.85) |
| Time from needle to puncture (median, IQR) | 35.5 (21 - 58) [n=14] | 52 (18 - 74) [n=10] | 0.78 (0.71 - 0.87) | 0.61 (0.54 - 0.69) |
| eTICI score of ≥2b on initial angiography of EVT (n, %) | 1 (7.14) | 0 (0.00) | - | - |
| Time from needle (IVT start) to first successful reperfusion (median, IQR) | 82.5 (44 - 133.5) [n=12] | 116 (49.5 - 150.5) [n=8] | 1.05 (0.96 - 1.13) | 0.80 (0.72 - 0.89) |
| Time from arterial access to successful reperfusion (median, IQR) | 29.5 (13 - 50.5) [n=12] | 23 (19 - 49) [n=8] | 1.2 (1.03 - 1.40) | 0.94 (0.78 - 1.12) |

Onset: stroke symptom onset. Door: hospital arrival. Needle: intravenous thrombolysis start. Puncture: groin puncture, EVT start. NIHSS, National Institutes of Health Stroke Scale. IQR: interquartile range.
Analysis: Poisson. Adjusted for age, sex, baseline NIHSS, intracranial occlusion, drug type and door-to-needle time.

**Figure 3.**
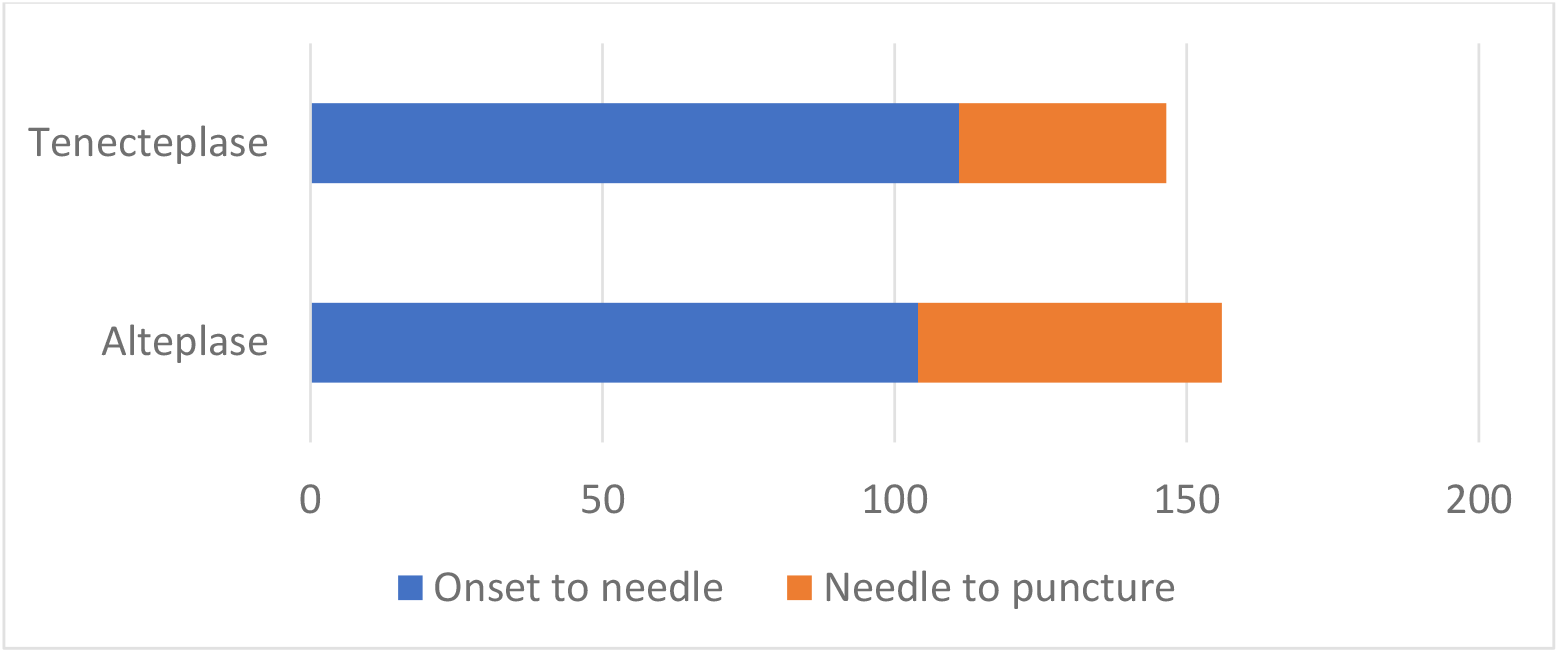
Onset-to-needle and needle-to-puncture times in patients transferred from PSCs to CSCs for potential EVT, by thrombolytic type. PSC: Primary Stroke Centers. CSC: Comprehensive Stroke Centers. Values correspond to median. Numbers represent minutes.

There was no significant interaction for primary and safety outcomes (Supplemental Table 5).

## Discussion

In this subgroup analysis of the AcT trial we found that patients treated with IVT at PSCs had higher rates of excellent functional outcome and return to baseline function with similar safety outcomes as compared to those who were treated at CSCs. Moreover, the workflow at PSCs was less efficient than CSCs, with slower door-to-needle, door-to-CT, door-to-puncture and door-to-reperfusion times. Interestingly, among patients who were treated in PSCs and subsequently transferred to CSCs for EVT, those who received tenecteplase had faster workflows, with more efficient CT-to-needle and needle-to-puncture times, compared to their counterparts who received alteplase.

It is possible that early arrival to a stroke center could account for improved functional outcomes at 90 to 120 days, since prompt IVT is a pivotal treatment in the hyperacute phase of the stroke, as is early clinical stabilization. While a well-defined “bundle of care” is described for hemorrhagic stroke,^10^ but not for patients with ischemic stroke, patients treated at PSCs may receive faster temperature, blood pressure, oxygenation and glycemic control than patients admitted to a CSC several minutes later in the very early phase of the ischemic stroke. Additionally, our unadjusted analysis showed that patients treated at PSCs stayed fewer days at hospital compared to patients at CSCs (a median of 3 versus 5 days, respectively). Further investigation is needed to explore whether the length of stay is associated with center type. For instance, this could involve examining whether the proximity of PSCs to patient’s families and communities contributes to shorter stays, or if shorter stays at PSCs reflect a less medically complex patient population.

Although it is well-known that one of the key predictors of length of stay is the severity of the stroke,^11^ these two groups could have unmeasured baseline clinical or sociodemographic differences that our registries did not capture, despite having similar age, NIHSS, EVT rate, hemorrhagic complications and serious adverse events.

The performance of PSCs and CSCs regarding time metrics and clinical outcomes has been explored previously. The Transfer to the Closest Local Stroke Center vs Direct Transfer to Endovascular Stroke Center of Acute Stroke Patients With Suspected Large Vessel Occlusion in the Catalan Territory, RACECAT trial, which enrolled patients with suspected LVO in a drip-and-ship versus mothership model, showed more patients receiving IVT at local stroke centers compared to EVT - capable stroke centers (60.4% and 47.5%, respectively), with a difference in onset-to-needle times (120 and 155 minutes, respectively), similar door-to-needle times (30 and 33 minutes, respectively) and similar clinical outcomes at 90 days.^12^ A cohort study with participants that received reperfusion therapies recruited from Get With the Guidelines Stroke registry, showed better door-to-needle times at CSCs and EVT-capable centers compared to PSCs, without significant difference in functional independence at discharge between CSCs and PSCs.^13^ Door-to-needle time is key time metric in acute stroke care. In patients treated with IVT with or without EVT, lower door-to-needle times led to better clinical outcomes including home time and overall mortality, with chances of good outcome improving with each 15-minute period of optimization in the door-to-needle time.^14^ Several simple and effective measures have been identified to improve this time metric, such as establishing a STAT stroke protocol, transporting patients to CT in the EMS stretcher and giving IVT near the CT room.^15^ In our prespecified sub analysis, despite prolonged door-to-needle and door-to-puncture times, a higher proportion of patients in PSCs had better clinical outcomes compared to CSCs. In the unadjusted analysis, onset-to-door time was lower in PSCs compared to CSCs (67 versus 83 minutes, respectively) likely due to geographic and logistical reasons.

Certain time metrics at PSCs differed depending on the thrombolytic that was used for patients transferred to CSCs. For instance, despite similar door-to-needle times among those receiving tenecteplase and alteplase (56.5 minutes for both), those who were transferred to a CSC for EVT and received tenecteplase presented with lower CT-to-puncture (60.5 versus 90.5 minutes) and needle-to-puncture (35.5 versus 52 minutes) times. Even though we did not collect door-in-to-door-out times, it is evident that the transportation to a CSC was more efficient in those who were treated with tenecteplase. The pharmacological advantages of tenecteplase compared to alteplase, its effectiveness, safety, and the evidence that supports its use are well described elsewhere.^16–19^ Additionally, there are several real-world experiences that have shown reassuring results when alteplase has been replaced by tenecteplase.^20–23^ The possibility to give a single bolus of tenecteplase, instead of a bolus plus a 1-hour of continuous administration of alteplase through an infusion pump, with the risk of lower plasma level if there are delays between bolus and infusion,^24^ explains its success in shortening key workflow times especially for patients that are transferred to a CSCs to receive EVT in ambulances without the capability of continuous infusion administration. We did not find differences in early recanalization between patients treated with tenecteplase or alteplase at PSCs. The EXTEND-IA

TNK trial (part 1), a randomized controlled trial of tenecteplase versus alteplase in patients with LVO treated in the 4.5 hour window, showed a substantial reperfusion of 22% and 10%, favoring tenecteplase.^25^ The sub-analysis of the AcT trial in patients with medium-vessel occlusions showed a significant difference in intracranial recanalization favoring tenecteplase,^26^ without difference in the subgroup analysis for patients with LVO.^27^

Our findings challenge the prevailing assumption that CSCs uniformly provide superior outcomes compared to PSCs. Specifically, the observation that patients who remained at PSCs demonstrated outcomes that were comparable, if not better, than those treated at CSCs raises important questions about the potential advantages of PSC care. These benefits may stem from highly standardized, guideline-adherent care pathways, more streamlined inpatient management, or faster transitions to rehabilitation services often integrated within PSC settings. The observed 13.5% absolute difference in outcomes between PSCs and CSCs is notable, approaching the effect size of thrombolysis itself. While this comparison is made cautiously, this ‘PSC effect’ highlights the importance of decentralized care models, particularly in geographically widespread or resource-limited regions, where equitable access to acute stroke care is critical. Future research should explore the mechanisms underlying this effect and assess whether specific organizational practices or resource allocations at PSCs contribute to these outcomes.

This study has certain limitations. First, the study population considers only patients enrolled in the AcT trial, thus generalizability to other regions and countries with different stroke systems of care may be limited. Second, the number of patients transferred to CSCs for potential EVT and its workflow characteristics are insufficient to draw firm conclusions regarding workflow and time metrics in this subgroup of patients. Third, this study is a subgroup analysis of a randomized controlled trial, thus it serves as a hypothesis-generating exploratory analysis. Finally, there may be patient selection factors impacting which patients remained at PSCs that were not captured by our collected metrics and could have affected results in this small sample.

## Acknowledgments

We thank the Canadian Stroke Consortium for their support of the trial recruitment efforts and the Canadian Institutes of Health Research (grant numbers 419722 and 450890), the Alberta Strategy for Patient Oriented Research Support Unit, Alberta Innovates, the Heart and Stroke Foundation, and the University of Calgary for funding support.

## Notes

### Competing Interest Statement

BKM has stock options in Circle NVI and has consulted for Biogen and Boehringer Ingelheim. LC has received payments by Servier and consulting fees from Ischaemavie RAPID, Circle NV, and Canadian Medical Protective Association. JS has a grant from Medtronic to the University of Manitoba. AMD has received consulting fees from Medtronic and honoraria from Boehringer Ingelheim. TTS has received consulting fees from Circle NVI. RHS has stock options in FollowMD and receives salary support for research from the Heart & Stroke Foundation of Canada, Sandra Black Centre for Brain Resilience & Recovery, and Ontario Brain Institute. AG reports consulting fees and honoraria from Alexion, Biogen Eisai, and Servier Canada; research support from Alberta Innovates, the Alzheimer Society of Canada, the Alzheimer Society of Alberta and Northwest Territories, Brain Canada, the Canadian Institutes of Health Research, Campus Alberta Neuroscience, the Government of Canada INOVAIT and New Frontiers in Research programs, the France-Canada Research Fund, the Heart and Stroke Foundation of Canada, Microvention, MSI Foundation, and Panmure House; and stock/stock options from SnapDx Inc and Collavidence Inc (Let?s Get Proof)? all outside the scope of the published work. All other authors declare no competing interests.

### Clinical Trial

NCT03889249

### Clinical Protocols

https://www.ahajournals.org/doi/10.1161/SVIN.122.000447

### Funding Statement

Canadian Institutes of Health Research, Alberta Strategy for Patient Oriented Research Support Unit.

### Author Declarations

The trial was regulated by Health Canada (Clinical Trials Application [CTA] number 231509) and by research ethics boards at participating centres.

